# Survey of Contaminated Percutaneous Injuries in Anesthesia Practitioners

**DOI:** 10.1101/2021.01.21.21249920

**Authors:** Reine Zbeidy, Joshua Livingstone, Vadim Shatz, Yehuda Raveh, Rofayda Gad, Ramona Nicolau-Raducu, Fouad G. Souki

## Abstract

**Background:** Anesthesia practitioners are at inherent risk for percutaneous injuries by blood-contaminated needles and sharp objects. These exposures may result in transmission of HIV and hepatitis viruses. Data about this occupational hazard from contaminated needles and sharp devices is limited and decades old. We conducted a web-based survey to assess the occurrence, reporting, characteristics, and outcome of contaminated percutaneous injuries (CPI) in anesthesia residents, fellows, and attendings.

**Methods:** After institutional research board approval, an email was sent to 217 anesthesia practitioners requesting their participation in an online survey about contaminated percutaneous injuries. Responses were collected from February through March 2020. Results are reported as absolute numbers and proportions with 95% confidence interval (CI).

**Results:** The overall survey response rate was 51% (110/217). 59% (65/110) (95% CI, 50–68) of participants reported having one or more contaminated percutaneous injury during their years of anesthesia practice (42% (21/50) of residents, 50% (4/8) of fellows, 77% (40/52) of anesthesia attendings). Prevalence of injuries related to attendings’ years of anesthesia practice was 69% (95% CI, 44–94) for 5-10 years, 62.5% (95% CI, 29–96) for 10-15 years, and 79% (95% CI, 63– 95) for greater than 15 years of practice.

35% (95% CI, 26–44) of participants reported having one or more CPI within the last 5 years (40% of residents, 50% of fellows, 29% of attendings). Occurrence of CPI within the last 5 years based on attending anesthesiologist years of practice was 57% for less than 5 years, 37.5% for 10-15 years, and 20% for 15-20 years of practice. 75% (95% CI, 65–85) reported the incident at the time of injury. 59% (95% CI, 48–70) of injuries were due to hollow bore needles. 50% (95% CI, 39–61) of total injuries were high risk. 26% of injured anesthesia practitioners received post-exposure prophylaxis and there were zero seroconversions.

**Conclusion:** Most anesthesiologists will sustain a contaminated percutaneous injury during their careers. Incidence of these injuries decreases with years of practice. Occurrence of these injuries is high among anesthesia residents, with the majority reporting their injuries. Half of the injuries are high risk with a quarter requiring postexposure prophylaxis. More education and interventions are needed to reduce percutaneous injuries and improve reporting.

## Introduction

Anesthesia practitioners are at occupational hazard for percutaneous injuries by blood-contaminated needles and sharp objects. These exposures may result in transmission of HIV and hepatitis viruses. The risk of infection varies with the pathogen and type of exposure (e.g., superficial or deep, solid bore or hollow-bore needle) [1]. Data about the risk to anesthesia residents, fellows, and attendings from contaminated needles and other sharp devices is limited and decades old [2-7]. These injuries are often under reported which further hinders accurate data collection [6,7]. We conducted a web-based survey to assess the occurrence, reporting, characteristics, and outcome of contaminated percutaneous injuries (CPI) in anesthesia residents, fellows, and attendings.

## Methods

We obtained institutional research board approval to conduct a web-based survey to assess the occurrence, reporting, characteristics, and outcome of contaminated percutaneous injuries (CPI) in anesthesia residents, fellows, and attendings at a multihospital based anesthesia practice in south Florida, USA. An email was sent to all registered anesthesia practitioners (217) requesting their participation in an online survey. Respondents were informed that participation in the survey implied consent. Anesthesia practitioners consisted of 85 residents, 17 fellows, and 115 attendings. The survey was confidential and anonymous. No personal identifiers were collected, and the survey responses could not be traced back to responders. Non-delivered email was not reported. Since we could not identify responders and non-responders, a second email request for participation in the survey was sent after 2 weeks. Responses were collected from February through March 2020. The web-based survey was developed using SurveyMonkey^R^ (Portland, OR) following guidelines to survey research in anesthesiology [8]. The survey was pilot tested with 10 anesthesiologists and adjustments were made based on their remarks. The survey consisted of nine close-response questions, and two open-response questions (Appendix). The closed-response questions inquired about age, gender, job title, years of anesthesia practice, number of contaminated percutaneous injuries, contaminated percutaneous injury within the last 5 years, injury reporting, postexposure prophylaxis, and seroconversion. In the two open-response questions, respondents were asked to “choose all that apply” and/or provide a free-text answer about the device that caused injury and the presumed infectious status of source. SurveyMonkey^R^ prevented duplication of responses by allowing only one response per electronic device behind the internet protocol address. Survey results were reported as absolute values and proportions with 95% confidence interval (CI). Statistical analysis was performed using the normal approximation method, Wald’s method, to calculate the 95% CI for proportions [9,10]. This manuscript adheres to the applicable EQUATOR guidelines (STROBE).

## Results

We received 110 responses out of 217 email invitations, a 51% response rate. Survey completion rate was 99%. 60% of respondents were males. Survey response rate was 59% (50/85) for residents, 47% (8/17) for fellows, and 45% (52/115) for attendings. In all, 59% (65/110) (95% CI, 50–68) reported being injured percutaneously by a contaminated sharp object or needle (42% (21/50) of residents, 50% (4/8) of fellows, 77% (40/52) of anesthesia attendings) (Table 1). 54% reported being injured once, 34% injured twice, 12% injured three or more times. Survey participants reported 113 CPIs. Numbers of CPIs per anesthesia practitioner who answered survey was 0.58 for residents, 0.75 for fellows, and 1.5 for attendings.

**Table 1.** Survey data.

|  | ALL | Residents | Fellows | Attendings |
| --- | --- | --- | --- | --- |
| Surveys sent | 217 |  |  |  |
| Surveys returned | 110/217<br>51% | 50/85<br>59% | 8/17<br>47% | 52/115<br>45% |
| Anesthesiologists with CPI | 65/110<br>59% | 21/50<br>42% | 4/8<br>50% | 40/52<br>77% |
| Anesthesiologist with 1 CPI | 35<br>54% | 14/21<br>67% | 2/4<br>50% | 19/40<br>47.5% |
| Anesthesiologist with 2 CPI | 22<br>34% | 6/21<br>28% | 2/4<br>50% | 14/40<br>35% |
| Anesthesiologist with $\geq 3$ CPI | 8<br>12% | 1/21<br>5% | 0 | 7/40<br>17.5% |
| Number of CPI per anesthesiologists who answered survey | 113/110<br>1.03 | 29/50<br>0.58 | 6/8<br>0.75 | 78/52<br>1.5 |
| Anesthesiologists with CPI last 5 years | 39/110<br>35% | 20/50<br>40% | 4/8<br>50% | 15/52<br>29% |
| Anesthesiologists reported CPI at time of injury | 48/64<br>75% | 17/20<br>85% | 4/4<br>100% | 27/40<br>67.5% |
| Anesthesiologists received postexposure prophylaxis | 17/64<br>26.6% | 6/20<br>30% | 3/4<br>75% | 8/40<br>20% |

Prevalence of injuries related to attendings’ years of anesthesia practice was 69% (95% CI, 44– 94) for 5-10 years, 62.5% (95% CI, 29–96) for 10-15yrs, and 79% (95% CI, 63–95) for >15 years.

29% (15/52) (95% CI, 17-41) of attendings reported being injured within last 5 years compared to 40% of residents and 50% of fellows. Analyzing relationship between CPI within last 5 years and attending anesthesiologist’s years of practice revealed an incidence of 57% for less than 5 years of practice, 31% for 5-10 years of practice, 37.5% for 10-15 years of practice, 20% for 15-20 years of practice, and 14% for more than 20 years of practice (Table 2). 75% (48/64) (95% CI, 64–86) reported the incident at the time of injury (85% of residents, 100% of fellows, 67.5% of attendings).

**Table 2.** Percutaneous injuries reported by attending anesthesiologists compared to years of practice (Total).

| Years of practice | <5 | 5-10 | 10-15 | 15-20 | >20 |
| --- | --- | --- | --- | --- | --- |
| Attendings responded (52) | 7<br>13.5% | 13<br>25% | 8<br>15.5% | 10<br>19% | 14<br>27% |
| Anesthesiologists reported CPI (40) | 7/7<br>100% | 9/13<br>69% | 5/8<br>62.5% | 8/10<br>80% | 11/14<br>78.5% |
| Number of CPI per anesthesiologists who answered survey (78) | 10/7<br>1.43 | 13/13<br>1 | 10/8<br>1.25 | 20/10<br>2 | 25/14<br>1.8 |
| Reported having CPI in last 5 years (15) | 4/7<br>57% | 4/13<br>31% | 3/8<br>37.5% | 2/10<br>20% | 2/14<br>14% |

59% (45/76) (95% CI, 48–70) of percutaneous injuries were due to hollow-bore objects and 41% were due to sharp objects (suture needle, scalpel, neuromonitoring needle) (Table 3). 84% (38/45) of hollow bore injuries or 50% (38/76) of all CPI were high risk (blood contaminated hollow-bore needle and lumen filled with undiluted blood).

**Table 3.** Devices causing contaminated percutaneous injuries (CPI).

|  | Type of Device | CPI (n) |
| --- | --- | --- |
| Solid<br>41% | Suture needle | 21 |
|  | Scalpel | 6 |
|  | Other (J tube wire, head frame pins) | 2 |
|  | Neuromonitoring needle | 2 |
| Hollow-bore<br>59% | Spinal or Epidural needle | 4 |
|  | Nerve block needle | 3 |
|  | Hollow-bore needle (venous, arterial, central) | 38 |
| Total |  | 76 |

At the time of CPI, 16% (10/61) knew that the source had a history of blood-borne infection (1 HBV, 4 HCV, 4 HIV, 1 HIV+HCV). When asked about postexposure prophylactic treatment, 26.6% (17/64) reported receiving it. 70.5% (12/17) of those who received HIV postexposure prophylactic treatment did not know, at the time of injury, that the source had an HIV infection. None (0/64) of those who sustained percutaneous injury seroconverted.

## Discussion

This study revealed how incidence of CPI changes with years of practice, demonstrated how reporting of injuries has improved, described the specific nature of injuries in anesthesia practitioners, and assessed the need for postexposure prophylactic treatment after CPI. Results revised previously published data about incidence, reporting, characteristics, and outcome of percutaneous injuries in anesthesia residents, fellows, and attendings.

### Rate of injury

Only few reports in the medical literature specifically address anesthesiologists’ risk of occupational exposure and infection after CPI [2-7]. These reports, dating back to the 90s, have estimated an overall percutaneous exposure incidence of 1.35 CPIs per 1000 anesthetics, 0.42 CPIs per year per full time equivalent, 0.54 CPIs per 1000 hours of anesthesia, and 0.27 CPIs per year per person [2-4,7,11]. However, these estimates did not consider the effect of anesthesiologists’ years of practice and experience on incidence of CPI. This study highlighted that most anesthesiologists will sustain a percutaneous injury during their careers, and that the incidence of CPI decreases with each 5 years of practice. (Table 2). Anesthesiologists with less than 15 years of practice had more than twice the incidence of CPI in the last 5 years compared to anesthesiologists with more than 15 years of practice (39% (11/28) versus 17% (4/24)). Other studies, not specific to anesthesiologists, also found that incidence of injuries gradually decreased with experience [12,13]. These findings underscore the importance of education and experience in the early years of anesthesia practice in preventing the occurrence of CPI thereafter.

Several studies have documented CPI in various hospital departments, but none have been specific to anesthesia residents or fellows [14-19]. Our data showed that almost half of anesthesia residents and fellows sustain a CPI. Similarly, a survey targeting residents in the operative setting revealed that 34% (11/32) of anesthesia residents had a CPI compared to 99% of general surgery residents by the end of training [14,15].

We did not inquire about incidence of CPI per postgraduate year of training or the circumstances of CPI in residents and fellows. However, surveys conducted in surgical residents showed that the mean number of needle stick injuries increases with every postgraduate year of training [15,16]. Lapses in concentration, fatigue, inexperience, feeling rushed, long work hours, and sleep deprivation have been attributed to CPIs [14,15,17-19]. Fatigue was reported as a contributing factor in 2 out of 3 of anesthesia residents compared to only 1 out of 5 surgical residents [14]. More research into causes of CPI in anesthesia residents and fellows is needed.

### Reporting

Timely reporting of occupational exposures to an employee health service is necessary to ensure appropriate counseling, facilitate prophylaxis or early treatment, and establish legal prerequisites for workers’ compensation [7, 14, 20]. Reporting can reduce rates of injury by identifying risk-prone behaviors and practices, and guide improvements in prevention [4]. Historically, reporting of needle sticks and sharp injuries to the workplace monitoring system by health care workers was 30% in the early 90s and averaged 50% in the late 90s and early 2000 [7, 15, 21-24]. In our survey, reporting of CPI by anesthesia attendings, residents, and fellows has tripled when compared to 19-29% historic numbers available in the anesthesia literature [7]. This denotes a significant improvement in reporting, although less than perfect. It seems that the same conditions that marred reporting of CPI still exist (perception of low risk, paperwork, time consuming, lack of knowledge about cost coverage) [14,15]. More education about the risks of disease transmission and emphasis on benefits of reporting injuries is needed to drive rates higher, particularly in anesthesia attendings [14,20].

### Type of injury

Anesthesiologists are unique compared to other nonanesthesia operating room personnel in that they are more likely to sustain percutaneous injuries from hollow-bore needles than from sharp objects [6, 15, 16, 25]. Blood-contaminated hollow-bore injuries carry a higher risk of pathogen transmission than injuries from sharp solid objects [6, 15, 16]. In the 90s, Greene reported that 74% of CPI in anesthesiologists were associated with hollow-bore needles, and 30% of all CPI were high risk [7]. These trends continued in our survey with some decrease in percentage of hollow-bore needle injuries and increase in percentage of high-risk injuries. This may be explained by increased compliance with work practice modifications (avoidance of hollow-bore needle recapping) and prevalence of engineering solutions (shielding of hollow-bore needle tips after use).

### Outcome

The risk of occupational infection with a blood-borne pathogen is proportional to the prevalence of patients carrying the pathogen, the risk of infection transmission, and the number of exposures to blood or body fluids [4, 6, 26]. In the hospital surgical setting, prevalence of HIV, HBV, and HCV is higher than that in the community [27]. Reports showed that 68.4% of seropositive cases for HBV, HCV, and HIV detected by preoperative screening tests were previously undiagnosed [15, 23]. Similarly, 70% of those who received HIV prophylaxis in our survey did not know that the source had HIV infection at the time of injury. This confirms that there is a much higher risk of exposure to blood-borne pathogens than can be evaluated based on medical history alone perioperatively [14].

Published data estimates the risk of infection transmission after accidental percutaneous exposure to contaminated blood of 0.3% for HIV, 2% for HCV, and up to 30% for HBV susceptible practitioners without post exposure prophylaxis or sufficient hepatitis B vaccination [13, 28-30]. However, two recent studies in which 21% of health care workers received postexposure HIV prophylaxis reported 0% (0/266) HIV and 0.1% (2/1361) HCV seroconversions after percutaneous and mucocutaneous exposures to infected blood and body fluids [31, 32]. Results of our survey were similar and emphasized the frequent need for postexposure prophylaxis (26% (17/64) after CPI and the very low seroconversion rates (0/64).

Since the number of exposures to blood or body fluids could be altered, it is essential that anesthesiologists know and practice strategies that minimize percutaneous injuries. To this end, anesthesiologists must receive adequate training and supervision, avoid recapping by the two-handed technique, appropriately use needle and syringe disposal containers, dress all abrasions and cuts, eliminate unnecessary sharp devices, use needleless or protected needle devices, and wear protective barriers (personal protective clothing, gloves, masks, face shields) [4,7,28,33,34]. In addition, anesthesiologists should be informed about the importance of reporting injuries and following up with testing and/or prophylactic treatment [29].

### Limitations

The results obtained are comparable to published literature and we have no reason to believe that its findings are not representative of the wider problem. However, some limitations should be considered. First, the survey is retrospective with likelihood of recall bias and overestimating or underestimating exposure incidents. Second, the response rate was 51% although the survey was short, anonymous, and a reminder to participate was sent. Such response rate underscores the possibility of non-response bias that could have selected those with strong attitudes towards the subject [35]. Non-response could also be due to nondelivered email, unfamiliarity with topic, and lack of motivation to participate. Nevertheless, some reports have called into question the possible association of nonresponse rate and response bias [36,37]. Third, the study is relatively small with possible concomitant attenuation of statistical power and precision [37]. In that regards, the maximum half width 95% CI for those who reported a percutaneous injury was 9%. A future direction is a larger study to produce more data and allow for a more detailed subgroup analysis.

In conclusion, most anesthesiologists will sustain a contaminated percutaneous injury during their careers. Incidence of these injuries decreases with years of practice. Occurrence of these injuries is high among anesthesia residents, with the majority reporting their injuries. Most CPI in anesthesia personnel are due to hollow-bore needles with half of CPI being high risk and one-fourth receiving postexposure HIV prophylaxis. These findings underscore the need for more education and interventions to reduce occupational blood exposures and improve reporting.

## Data Availability

All the data is included in the manuscript.

## Appendix

### Survey Questions

1. What is your Age?
  A. 25-34
  B. 35-44
  C. 45-54
  D. 55-64
  E. > 65
2. What is your gender?
  A. Female
  B. Male
3. Which of the following best describes your current job level?
  A. Resident
  B. Fellow
  C. Attending
4. About how many years have you been practicing anesthesia?
  A. Less than 5 years
  B. At least 5 years but less than 10 years
  C. At least 10 years but less than 15 years
  D. At least 15 years but less than 20 years
  E. 20 years or more
5. In your years of anesthesia practice, how many times have you been injured by a CONTAMINATED sharp object/needle?
  A. 0
  B. 1
  C. 2
  D. 3
  E. 4
  F. 5
  G. More than 5
6. Have you had any accidental injury by a CONTAMINATED sharp object/needle in the past 5 years?
  A. Yes
  B. No
7. Which of the following contaminated devices caused the injury? (Choose all that apply)
  A. Suture needle
  B. Regional anesthesia needle
  C. Spinal or epidural needle
  D. Hollow-bore needle (venous, arterial, central)
  E. Sharp object (scalpel)
  F. Other (please specify)
8. What was the viral status of the patient at the time of injury? (Choose all that apply)
  A. HIV positive
  B. HBV positive
  C. HCV positive
  D. Unknown
  E. Other (please specify)
9. Did you report your injury to your supervisor or medical health services when it occurred?
  A. Yes
  B. No
10. Did you receive post exposure prophylactic treatment?
  A. Yes
  B. No
11. Did you seroconvert (i.e. acquire infection)?
  A. Yes
  B. No

